## Supplementary Files for "The role of the microbiota in respiratory virus-bacterial pathobiont relationships in the upper respiratory tract"

**SUPPLEMENTARY INFORMATION**

| **Supplementary Table 1.** Associations between respiratory virus infections and acute respiratory infection symptoms | | | | | |
| --- | --- | --- | --- | --- | --- |
|  | | **Sample prevalence** | | **ARI symptoms** | |
|  | | **n** | **(%)** | **n** | **(%)** |
| Adenovirus | | 65 | 2.7% | 23 | 35% |
| Human metapneumovirus | | 17 | 0.7% | 13 | 76% |
| Influenza viruses | | 14 | 0.6% | 11 | 79% |
|  | A | 13 | 0.5% | 11 | 85% |
|  | B | 1 | <0.1% | 0 | 0% |
| Parainfluenza viruses | | 39 | 1.6% | 23 | 59% |
|  | Type 1 | 8 | 0.4% | 2 | 25% |
|  | Type 2 | 4 | 0.2% | 1 | 25% |
|  | Type 3 | 27 | 1.1% | 20 | 74% |
| Rhinovirus/enterovirus | | 507 | 21.4% | 209 | 41% |
| Respiratory syncytial virus | | 44 | 1.9% | 32 | 73% |
| SARS-CoV-2 | | 1 | <0.1% | 0 | 0% |
| More than one virus | | 48 | 2.0% | 23 | 48% |
| No viruses | | 1733 | 73.1% | 308 | 18% |
| ARI, acute respiratory infection; SARS-CoV-2, severe acute respiratory syndrome coronavirus 2 | | | | | |

| **Supplementary Table 2.** Associations between respiratory virus infections and the odds of acquisition of bacterial pathobionts during infancy | | | | | | | | |
| --- | --- | --- | --- | --- | --- | --- | --- | --- |
|  | | | **Intervals**  **(n)** | **Colonization frequency**  **(n, %)** | | **Odds ratio** | **95% confidence interval** | **p** |
| *H. influenzae* acquisition | | |  |  |  |  |  |  |
|  | Any respiratory virus | | 431 | 110 | 26% | 1.44 | (1.06-1.96) | 0.02 |
|  |  | Rhinovirus/enterovirus only | 324 | 79 | 24% | 1.38 | (0.98-1.94) | 0.07 |
|  |  | Other respiratory viruses | 107 | 31 | 29% | 1.65 | (0.98-2.76) | 0.06 |
|  | No viruses detected | | 1081 | 180 | 17% | Ref | - | - |
| *M. catarrhalis* acquisition | | |  |  |  |  |  |  |
|  | Any respiratory virus | | 242 | 109 | 45% | 1.34 | (0.96-1.87) | 0.09 |
|  |  | Rhinovirus/enterovirus only | 187 | 79 | 42% | 1.22 | (0.84-1.77) | 0.29 |
|  |  | Other respiratory viruses | 55 | 30 | 55% | 1.82 | (0.96-3.45) | 0.06 |
|  | No viruses detected | | 729 | 256 | 35% | Ref | - | - |
| *S. aureus* acquisition | | |  |  |  |  |  |  |
|  | Any respiratory virus | | 512 | 30 | 6% | 0.45 | (0.29-0.70) | 0.0004 |
|  |  | Rhinovirus/enterovirus only | 382 | 24 | 6% | 0.46 | (0.28-0.75) | 0.002 |
|  |  | Other respiratory viruses | 130 | 6 | 5% | 0.39 | (0.15-1.00) | 0.0499 |
|  | No viruses detected | | 1177 | 170 | 14% | Ref | - | - |
| *S. pneumoniae* acquisition | | |  |  |  |  |  |  |
|  | Any respiratory virus | | 284 | 120 | 42% | 1.83 | (1.33-2.52) | 0.002 |
|  |  | Rhinovirus/enterovirus only | 219 | 92 | 42% | 1.87 | (1.32-2.64) | 0.0005 |
|  |  | Other respiratory viruses | 65 | 28 | 43% | 1.73 | (0.95-3.13) | 0.07 |
|  | No viruses detected | | 810 | 212 | 26% | Ref | - | - |
| Odds ratios, 95% confidence intervals, and p values were estimated from mixed effect logistic regression models adjusted for age, sex, low birth weight (<2500g), maternal HIV infection, location of residence, number of children under 5 years of age in the household, season, breastfeeding, receipt of antibiotics since the prior study visit, and 13-valent pneumococcal conjugate vaccine doses (for *S. pneumoniae* only). Ref, reference | | | | | | | | |

| **Supplementary Table 3.** Targets for multiplex PCR assay for the detection of bacterial pathobionts | |
| --- | --- |
| **Species** | **Primer and probe sequences** |
| *Haemophilus influenzae* | F, 5’ d CCAATCACATCATCTGCAACTA 3’  R, 5’ d GCGTGGCTTGATGGATTG 3’  Probe, 5’ d CAL Fluor Red 610-TGGTTTTTCGCCTGACTCACTAAAAG-BHQ-2 3’ |
| *Moraxella catarrhalis* | F, 5’ d GGTAAAAAGGTTGGAAAAACC 3’  R, 5’ d CTGACGCTAAGATTAGGATTGA 3’  Probe, 5’ d Quasar 670-CCAAGCTTTGGGGTGATTTATGATG-BHQ-2 3’ |
| *Staphylococcus aureus* | F, 5’ d TGMATATTTTACCTAGTAAAGAAGG 3’  R, 5’ d ATACRTACATTCAAATCAGCTTC 3’  Probe, 5’ d HEX-TGTACGAAGAATTRGAAAGAACGATTGAG-BHQ-1 3’ |
| *Streptococcus pneumoniae* | F, 5’ d AAGACTTCAATAAGCCTTCTACG 3’  R, 5’ d GAGAAGGTGAGRAGTCAAGAAG 3’  Probe, 5’d FAM-TCTTCAGACGAATTTTCTGTCATAGGAACA-BHQ-1 3’ |

**Supplementary Figure 1. Comparison of upper respiratory tract microbiota types identified using k-medoids and k-means clustering. a.** NMDS plot depicts microbiota types identified using k-medoids clustering on Bray-Curtis distances. Ellipses define the regions containing 80% of all samples that can be drawn from the underlying multivariate t distribution. **b.** Relative abundances of highly abundant genera in infant nasopharyngeal samples by k-medoids microbiota type. **c.** Classification of microbiota types using k-means clustering on Euclidean distances by k-medoids microbiota type. **d.** NMDS plot depicts microbiota types identified using k-means clustering on Euclidean distances. Ellipses define the regions containing 80% of all samples that can be drawn from the underlying multivariate t distribution. **e.** Relative abundances of highly abundant genera in infant nasopharyngeal samples by k-means microbiota type. The composition of the microbiota types identified by k-means clustering is similar to the composition of the types identified by k-medoids clustering. **f.** Relative abundances of microbiota types identified using k-medoids clustering on Bray-Curtis distances by k-means microbiota type. Overall, 1,943 of 2,235 (87%) samples with URT microbiota data had the same microbiota type classification using k-medoids or k-means clustering. URT microbiota types are abbreviated CD, *Corynebacterium/Dolosigranulum*-dominant; CDM, *Corynebacterium/Dolosigranulum/Moraxella*-dominant; *COR*, *Corynebacterium*-dominant; HAE, *Haemophilus*-dominant; MOR, *Moraxella*-dominant; STA, *Staphylococcus*-dominant; STR, *Streptococcus*-dominant; and OTH, “other.” ASV, amplicon sequence variant; NMDS, non-metric multidimensional scaling

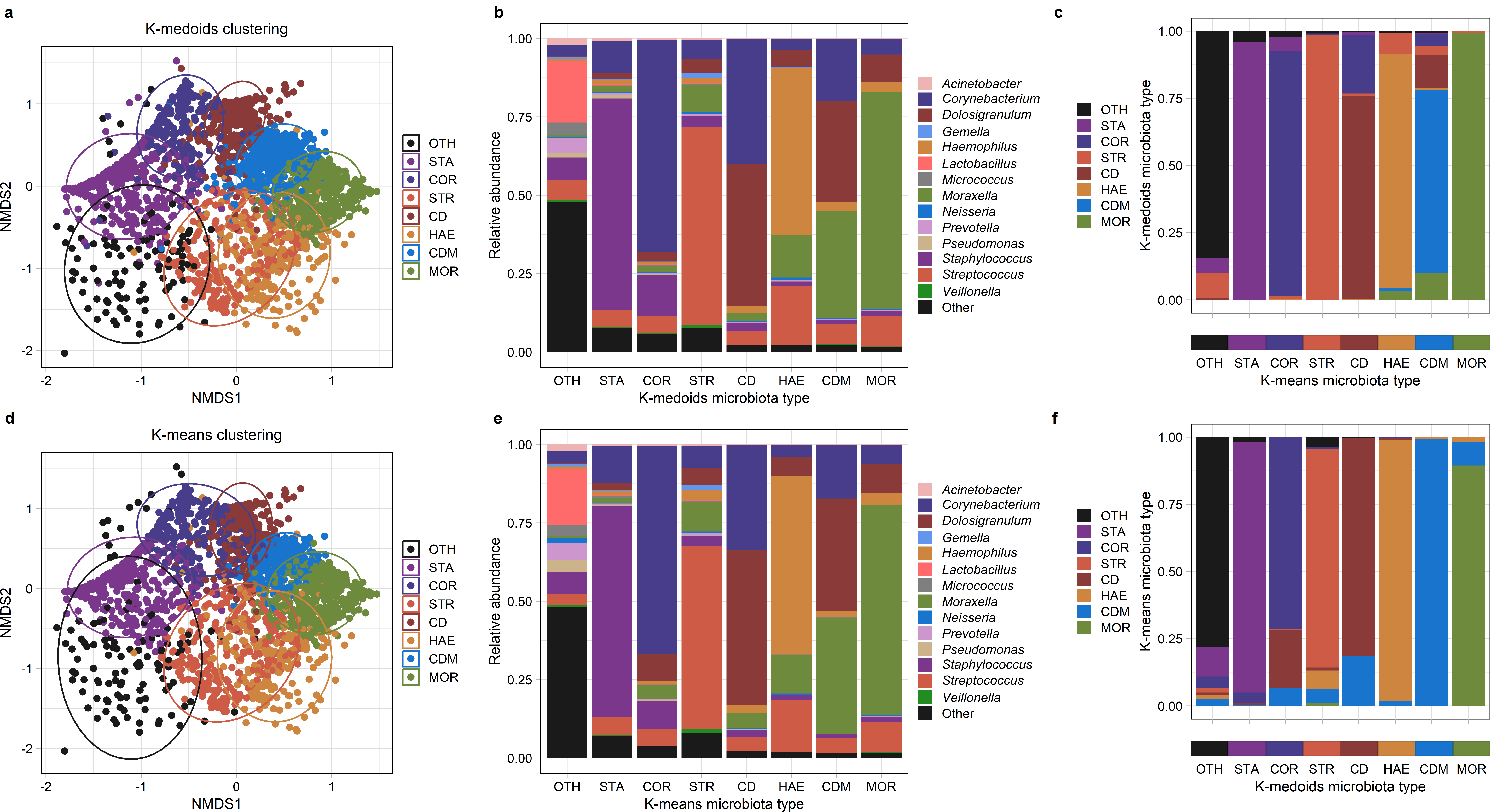

**Supplementary Figure 2. Shifts in upper respiratory microbiota community states associated with the acquisition of bacterial pathobionts.** Alluvial diagrams depict transitions from microbiota types associated with the acquisition of (**a.**) *H. influenzae*, (**b.**) *M. catarrhalis*, (**c.**) *S. aureus*, and (**d.**) *S. pneumoniae*. Microbiota types dominated by the genus corresponding to the bacterial pathobiont are opaque, while other microbiota types are depicted as semi-transparent. URT microbiota types are abbreviated CD, *Corynebacterium/Dolosigranulum*-dominant; CDM, *Corynebacterium/Dolosigranulum/Moraxella*-dominant; *COR*, *Corynebacterium*-dominant; HAE, *Haemophilus*-dominant; MOR, *Moraxella*-dominant; STA, *Staphylococcus*-dominant; STR, *Streptococcus*-dominant; and OTH, “other.”

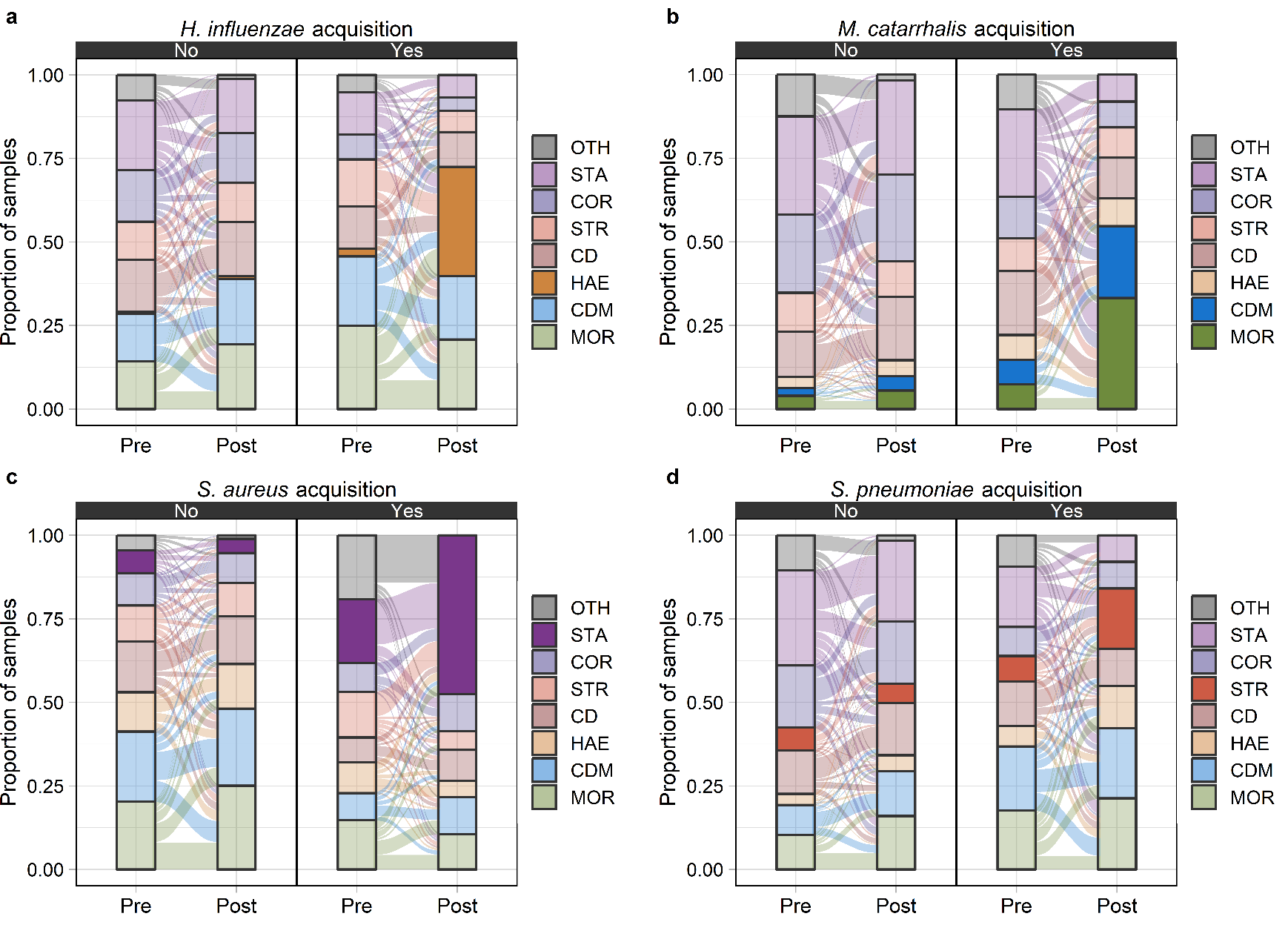

**Supplementary Figure 3. Identification of ASVs containing bacterial respiratory pathobionts. a.** Violin and box plots depict the relative abundances of specific ASVs within the upper respiratory microbiota by pathobiont colonization status. Based on BLAST searches, higher relative abundances of several ASVs potentially containing a pathobiont were observed among children identified as being colonized with that pathobiont by PCR, suggesting that these ASVs contain that pathobiont species. Specifically, higher relative abundances of ASVs 7 and 8 were observed with *H. influenzae* colonization, higher relative abundances of ASV1 were observed with *M. catarrhalis* colonization, higher relative abundances of ASV3 were observed with *S. aureus* colonization, and higher relative abundances of ASV5 were observed with *S. pneumoniae* colonization. In contrast, lower relative abundances of ASV15 were observed in children with *S. pneumoniae* colonization, consistent with this ASV being classified as other streptococcal species using BLAST. Wilcoxon rank-sum tests were used to estimate p values for differences in ASV relative abundances by pathobiont colonization status. **b.** Scatterplots show the correlation between log-transformed pneumococcal colonization density and the relative abundances of specific ASVs within the upper respiratory microbiota. Spearman’s rank correlation coefficients and associated p value are depicted. A strong positive correlation (ρ=0.83) was observed between the pneumococcal colonization density by PCR and the relative abundance of ASV5, providing further evidence that this ASV contains the pathobiont *S. pneumoniae*. A weak negative correlation (ρ=-0.22) was observed between the colonization density and the relative abundance of ASV15, consistent with this ASV not containing *S. pneumoniae*. ASV, amplicon sequence variant

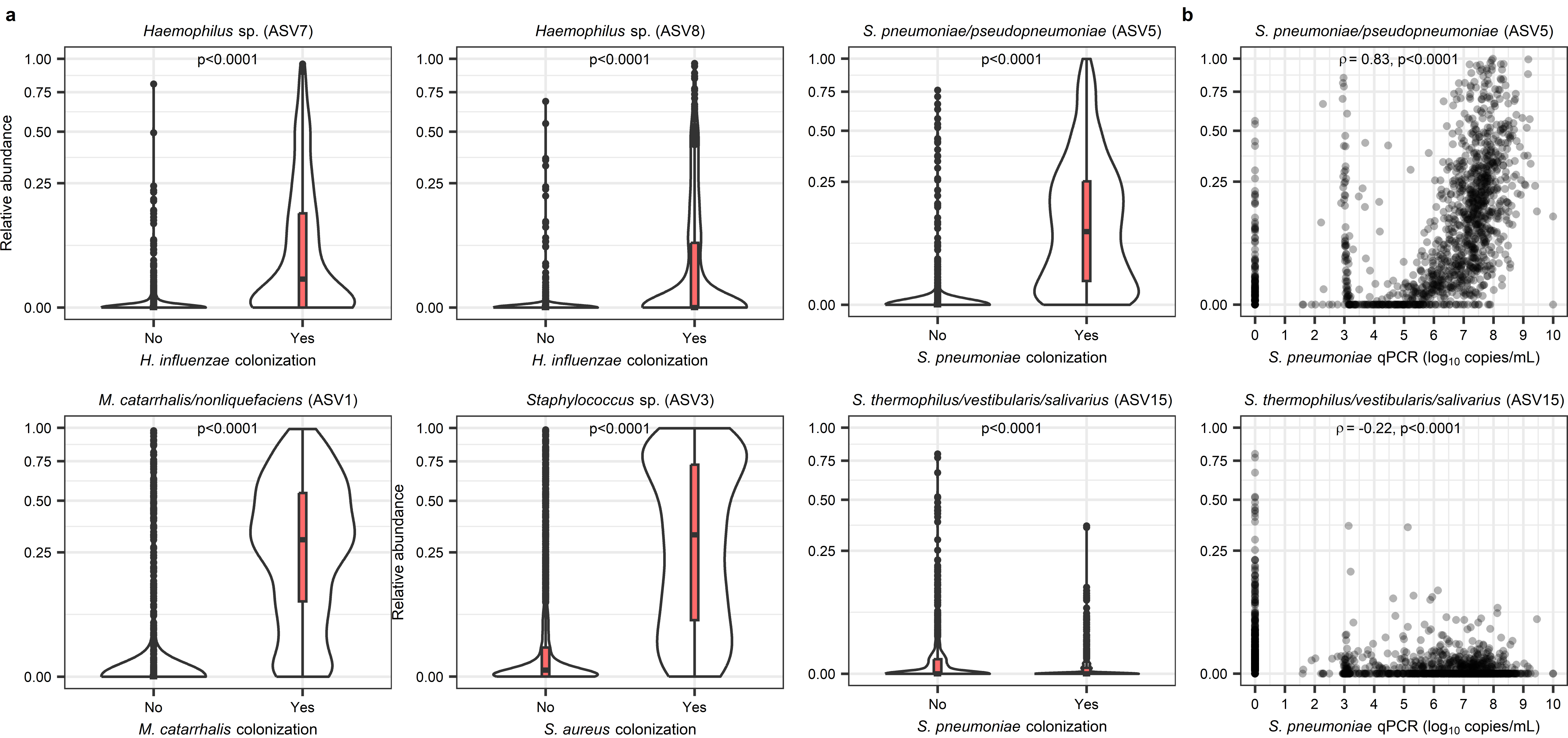
